# Epidemiological feature, viral shedding, and antibody seroconversion among asymptomatic SARS-CoV-2 carriers and symptomatic/presymptomatic COVID-19 patients

**DOI:** 10.1101/2020.12.18.20248447

**Authors:** Yi Chen, Ping Li, Yi-Bo Ding, Miao Liu, Lei-Jie Liu, Bo Yi, Ting Wu, Hong-Jun Dong, Xu-Ying Lao, Ke-Qing Ding, Hai-Bo Wang, Dong-Liang Zhang, Xiao-Jie Tan, Zhong-Fa Wang, Guo-Zhang Xu, Guang-Wen Cao

## Abstract

Novel coronavirus disease 2019 (COVID-19) caused by severe acute respiratory syndrome coronavirus 2 (SARS-CoV-2) is pandemic. However, data concerning the epidemiological features, viral shedding, and antibody dynamics between asymptomatic SARS-CoV-2 carriers and COVID-19 patients remain controversial. We enrolled 193 subjects infected with SARS-CoV-2 in Ningbo and Zhoushan, Zhejiang, China from January 21 to March 6, 2020. All subjects were followed up to monitor the dynamics of immunoglobulin M (IgM) and IgG against SARS-CoV-2. Of those, 31 were asymptomatic carriers, 149 were symptomatic patients, and 14 were presymptomatic patients. Compared to symptomatic patients, asymptomatic carriers were younger and had higher levels of white blood cell and lymphocyte, lower levels of C-reactive protein and viral load, and shorter viral shedding duration. Conversion of IgM from positive to negative was shorter in asymptomatic carriers than in COVID-19 patients (*P*=0.030). The proportion of those persistently seropositive for IgG was higher in COVID-19 patients than in asymptomatic carriers (*P*=0.037). Viral load was higher in symptomatic than presymptomatic patients. Viral shedding was longer in presymptomatic patients than in asymptomatic carriers. Conclusively, asymptomatic carriers have a higher antiviral immunity to clear SARS-CoV-2 than do symptomatic patients and this antiviral immunity is not contributable to humoral immunity.

## INTRODUCTION

Novel coronavirus disease in 2019 (COVID-19) caused by severe acute respiratory syndrome coronavirus 2 (SARS-CoV-2) has being pandemic since it was firstly recognized in China in late December 2019 [1]. The case number keeps increasing. Globally, as of 9:42am CET, December 11, 2020, there have been 68,845,368 confirmed cases of COVID-19, including 1,570,304 deaths, reported to World Health Organization [2]. Family clustering and hospital-based transmission were the two major epidemiological features of this outbreak at the early stage, and continued to be an important cause of community-based SARS-CoV-2 transmission in a low prevalence region [3-6]. COVID-19 patients and asymptomatic carriers are the main sources of SARS-CoV-2 infection but might have differences in some features [7]. However, data concerning SARS-CoV-2 transmission and viral shedding duration between COVID-19 patients and asymptomatic SARS-CoV-2 carriers remain controversial. It has been summarized from early studies that viral load of asymptomatic carriers is comparable to symptomatic patients [8]. In another study, it has been demonstrated that a considerably higher viral load is present in samples from fatal cases compared to asymptomatic carriers [9]. Difference in the dynamics of antibody against SARS-CoV-2 between asymptomatic carriers and COVID-19 patients remains unknown. To provide the information for recognizing differences between asymptomatic and symptomatic SARS-CoV-2 infected subjects, we conducted a study to investigate the epidemiological feature, laboratory findings, viral shedding, and antibody conversion of SARS-CoV-2 infected cases among asymptomatic SARS-CoV-2 carriers and symptomatic/presymptomatic COVID patients in two cities, a low prevalence region in Zhejiang, China.

## METHODS

### Study design and patients

It is an ambispective cohort study. The study enrolled all confirmed cases with SARS-CoV-2 infection on Ningbo city and a familial clustering infection with SARS-Co-2 in Putuo district of Zhoushan city from January 21 to March 6, 2020. Epidemiological, clinical characteristics, pathogen and serological test results were collected by Ningbo Municipal Center for Disease Control and Prevention (Ningbo CDC) and CDC of Putuo district, Zhoushan (Putuo CDC). Some baseline information including demographic and pathogenic data of those patients were reported [10, 11]. After that, we continued to investigate viral load and viral shedding of SARS-CoV-2, laboratory tests, and the dynamics of serum immunoglobulin M (IgM) and immunoglobulin G (IgG) against SARS-CoV-2 between asymptomatic carriers and COVID-19 patients. All patients were followed up to monitor the dynamics of IgM and IgG against SARS-CoV-2. Those with antibody tests for two times or more within 160 days were included in antibody seroconversion analysis. Diagnoses and disease staging of COVID-19 were carried out according to the Protocol for the Diagnosis and Treatment of COVID-19 (Version 7th), National Health Commission of the People’s Republic of China [12]. Specifically, COVID-19 was diagnosed if the patient was tested positive for SARS-CoV-2 genomic RNA and accompanied by clinical symptoms including fever and cough. Asymptomatic carrier was identified in close contactors of COVID-19 patients if they did not have any symptom. We also classified as COVID-19 patients with onset of disease and those during the incubation period (presymptomatic). All diagnosed COVID-19 patients were classified as mild, common, severe, and extremely severe types. This study was approved by the Ethics Commissions of Ningbo CDC and Putuo CDC. Written informed consent was waived for emerging infectious diseases.

### Epidemiological survey

A semi-structured questionnaire was applied to obtain demographic information, exposure information of the familiar clustering cases via face-to-face interview and telephone calls by well-trained professionals. The data regarding any travel history to high risk areas with COVID-19 epidemic, contact with confirmed cases or asymptomatic carriers tested positive for SARS-CoV-2 genomic RNA, contact with patients with some symptoms like fever, dry cough, and expectoration in the past 2-3 weeks before illness onset. Any chance and duration of attending any kinds of population gatherings were recorded as well.

Clinical information included the date of symptom onset and admission to designated hospitals, clinical manifestation, routine laboratory examinations, and radiographic examinations. The clinical manifestations, chest computed tomography images, and laboratory results of patients in Ningbo were collected from the electronic medical record systems in the two designated hospitals: Ningbo First Hospital and Huamei Hospital. The information of patients from Zhoushan was collected from Zhoushan Maternal and Child Health Care Hospital and Putuo Hospital. Two researchers (PL and YD) independently reviewed all of the data to doubly check the accuracy of data collected.

### Examination of SARS-CoV-2 genomic RNA

Quantitative reverse transcription-PCR (qRT-PCR) assay was applied to detect SARS-CoV-2 genomic RNA in nasal and throat swabs, sputum, and feces of patients. Patients in Ningbo were examined using the test kits manufactured by Shanghai BioGerm Medical Technology (Shanghai, China) and Daan Gene Co., Ltd (Guangzhou, China) [13]. Patients in Zhoushan were examined using the test kits manufactured by Shanghai GeneoDx Biotech Company (Shanghai, China) [14]. Sample was positive if the cycling threshold (CT) values of reverse-transcription polymerase chain reaction (RT-PCR) for the ORF1ab and the N genes were less than 37. Sample was negative if no CT value, or CT value of greater than 40, or unrepeatable CT value in the range of 37-40.

### Detection of antibodies against SARS-COV-2

IgM and IgG against SARS-CoV-2 in the frozen reserved fasting serum samples were detected using the diagnostic enzyme-linked immunosorbent assay (ELISA) kits. The diagnostic kits used for Ningbo patients and Zhoushan patients were manufactured by Innovita Biological Technology (Tangshan, China) and Nanjing Wending Biotech Company (Nanjing, China), respectively [15]. Briefly, testing results by Innovita ELISA kits were determined by the color reaction. Dark band was considered positive. Light band indicated weak positive. Disappearance of the expected band was considered negative. Testing results by Wending ELISA kits were presented as OD/CO. Sample was positive if the ratio of optical density (OD) to the cut-off value (CO) was equal or greater than 1 and negative if the ratio was less than 1. A higher OD/CO value indicated a higher level of antibody concentration.

### Statistical Analysis

Categorical variables were presented as count (%) and compared using the χ^2^ test or the Fisher exact test. Continuous variables were described using median and interquartile range (IQR) values and then compared using Mann-Whitney U test or Kruskal Wallis test. These statistical analyses were two-sided and performed using R, version 3.6.2 (R Foundation for Statistical Computing, Canberra, Austria). Scatter diagram to demonstrate the distribution of IgM and IgG against SARS-CoV-2 among asymptomatic carriers and symptomatic and presymptomatic COVID-19 patients were generated by R software. A *P* value of <0.05 was considered significant for two independent groups. An adjusted *P* value of <0.017 was considered significant by Bonferroni-Dunn test for pairwise comparison among three groups.

## RESULTS

### Epidemiological characteristics of asymptomatic SARS-CoV-2 carriers, symptomatic COVID-19 patients, and presymptomatic COVID-19 patients

A total of 193 SARS-CoV-2 infected subjects were enrolled in this study. Of those, 31 were asymptomatic carriers, 148 were symtomatic COVID-19 patients, and 14 were presymptomatic COVID-19 patients. Of the 187 patients from Ningbo, 3 family clusters were included. Those patients were close contacts of confirmed COVID-19 patients and then tested positive for SARS-CoV-2 genomic RNA, from January 21 to March 6, 2020. For a family cluster from Putuo, a 41-year-old man, who once contacted with a COVID-19 relative, was the first case diagnosed as COVID-19. The other five family members were close contacts of the man. All family members were tested positive for SARS-CoV-2 genomic RNA, with 1 asymptomatic SARS-CoV-2 carrier and 5 symptomatic COVID-19 cases. Our epidemiological survey indicated the family members did not have opportunity to get the infection from other sources. Asymptomatic SARS-CoV-2 carriers were significantly younger than symptomatic COVID-19 patients and presymptomatic COVID-19 patients. Compared to symptomatic COVID-19 patients, asymptomatic SARS-CoV-2 carriers had higher levels of circulating white blood cell (WBC) and lymphocyte, lower levels of C-reactive protein (CRP) and viral load, and shorter viral shedding time. Interestingly, viral load was significantly lower in presymptomatic COVID-19 patients than in symptomatic COVID-19 patients. The viral shedding duration was significantly longer in presymptomatic COVID-19 patients than in asymptomatic carriers. The first-time serological tests showed that nearly one-third of asymptomatic carriers and symptomatic COVID-19 patients were seronegative for IgM against SARS-CoV-2 while the seronegative rates for IgG to SARS-CoV-2 were around 7% in the two populations, respectively (Table 1).

**Table 1.** Baseline information of COVID-19 patients and asymptomatic SARS-CoV-2 carriers in Ningbo and Zhoushan cities of Zhejiang province, China

|  | Asymptomatic<br>carriers<br>(N=31) | Symptomatic<br>COVID-19<br>patients<br>(N=148) | Presymptomatic<br>COVID-19<br>patients<br>(N=14) | P<sup>*</sup> | P<sup>&</sup> | P<sup>#</sup> |
| --- | --- | --- | --- | --- | --- | --- |
| Gender |  |  |  | 0.453 | 1.000 | 0.750 |
| Male | 15 (48.4) | 50 (33.8) | 7 (50.0) |  |  |  |
| Female | 16 (51.6) | 98 (66.2) | 7 (50.0) |  |  |  |
| Age, years | 42.00(24.00-55.00) | 53.00(38.00-62.75) | 55.00(39.50-71.00) | **<0.001** | **0.002** | 1.000<sup>§</sup> |
| <30 | 9 (29.0) | 17 (11.5) | 1 (7.1) |  |  |  |
| 30-59 | 18 (58.1) | 89 (60.1) | 8 (57.1) | 0.076 | 0.345 | 1.000 |
| ≥60 | 4 (12.9) | 42 (28.4) | 5 (35.7) |  |  |  |
| Underlying diseases |  |  |  | 1.000 | 1.000 | 1.000 |
| No | 24 (77.4) | 104 (71.7) | 9 (64.3) |  |  |  |
| Yes | 7 (22.6) | 41 (28.3) | 5 (35.7) |  |  |  |
| WBC, 10<sup>9</sup>/L | 5.83(5.00-7.11) | 4.63(3.80-5.69) | 5.83(4.73-6.75) | **<0.001** | 1.000 | 0.075<sup>§</sup> |
| Lymphocyte, 10<sup>9</sup>/L | 1.53(1.32-2.11) | 1.22(0.86-1.60) | 1.26(0.89-1.86) | **0.003** | 0.420 | 1.000<sup>§</sup> |
| CRP, mg/L | 1.00(0.60-2.99) | 6.90(2.04-18.20) | 3.07(0.97-14.45) | **<0.001** | 0.317 | 0.885<sup>§</sup> |
| Viral shedding, day | 24.00(21.00-30.80) | 46.50(35.00-58.00) | 48.00(23.75-51.25) | **<0.001** | **0.002** | 1.000<sup>§</sup> |
| qRT-PCR-CT values | 31.40(27.50-34.50) | 29.00(24.25-32.00) | 33.50(28.25-36.25) | **0.004** | 0.410 | **0.003**<sup>§</sup> |
| IgM |  |  |  | 1.000 | 1.000 | 1.000 |
| Negative | 12 (38.7) | 43 (31.9) | 5 (50.0) |  |  |  |
| Positive | 19 (61.3) | 91 (67.4) | 5 (50.0) |  |  |  |
| Weak Positive | 0 (0.0) | 1 (0.7) | 0 (0.0) |  |  |  |
| IgG |  |  |  | 0.112 | 1.000 | 0.504 |
| Negative | 2 (6.5) | 10 (7.4) | 1 (10.0) |  |  |  |
| Positive | 25 (80.6) | 122 (90.4) | 8 (80.0) |  |  |  |
| Weak Positive | 4 (12.9) | 3 (2.2) | 1 (10.0) |  |  |  |

Abbreviations: WBC, white blood cell; CRP, C-reactive protein; CT, cycling threshold; IQR, interquartile range.
P<sup>\*</sup>, Asymptomatic carrier vs. Symptomatic COVID-19 patients.
P<sup>&</sup>, Asymptomatic carrier vs. Presymptomatic COVID-19 patients.
P<sup>#</sup>, Symptomatic COVID-19 patients vs. Presymptomatic COVID-19 patients.
§Continuous variables are expressed as median (IQR). P value was calculated using Kruskal Wallis test.
Categorical variables are expressed as n (%). P values were calculated using $\chi^2$ test and Fisher's exact test.

### Dynamics in seroconversion of IgM and IgG against SARS-CoV-2 between asymptomatic carriers and COVID-19 patients

Of the 193 study subjects, 74 (15 asymptomatic carriers and 59 COVID-19 patients) had two consecutive test results of IgM and IgG against SARS-CoV-2 within 160 days. SARS-CoV-2-specific IgM seroconversion from positive to negative or weak positive occurred in 9 (60.0%) asymptomatic carriers, while this occurred in 28 (47.5%) COVID-19 patients (*P*=0.647). However, the median time interval of IgM seroconversion from positive to negative was 7.50 (IQR, 4.75-11.50) days in asymptomatic carriers, which was significantly shorter than 25.50 (IQR, 6.75-56.75) days in COVID-19 patients (*P*=0.030). SARS-CoV-2-specific IgG seroconversion from positive to negative or weak positive occurred in 8 (53.4%) asymptomatic carriers, while this occurred in 15 (25.5%) COVID-19 patients (*P*=0.059). Importantly, 5 (33.3%) of asymptomatic carriers were consistently seropositive for IgG against SARS-CoV-2, however, this was 39 (66.1%) in COVID-19 patients (*P*=0.037). Furthermore, there was no significant difference in the time interval of IgG seroconversion between the two groups (Table 2, Figure 1).

**Table 2.** Seroconversion of IgG and/or IgM antibody against SARS-CoV-2 during follow-up time

|  | Overall | Asymptomatic carrier<br>(N=15) | COVID-19 patients<br>(N=59) | *P* |
| --- | --- | --- | --- | --- |
| Seroconversion of IgM antibody |  |  |  | 0.647 |
| From positive to negative | 28(37.8) | 8(53.3) | 20(33.9) | 0.234 |
| From positive to weak positive | 9(12.2) | 1(6.7) | 8(13.6) | 0.676 |
| Consistent negative | 28(37.8) | 5(33.3) | 23(39.0) | 0.772 |
| Consistent positive | 9(12.2) | 1(6.7) | 8(13.6) | 0.676 |
| Time interval of IgM antibody seroconversion, days |  |  |  |  |
| From positive to negative | 13.00(6.00-33.75) | 7.50(4.75-11.50) | 25.50(6.75-56.75) | **0.030** |
| From positive to weak positive | 16.00(3.00-28.00) | 12.00(12.00-12.00) | 19.00(3.00-30.00) | 0.694 |
| Consistent negative | 11.50(7.00-25.50) | 27.00(10.00-37.00) | 11.00(7.00-22.50) | 0.186 |
| Consistent positive | 9.00(8.00-13.00) | 13.00(13.00-13.00) | 8.50(7.50-14.50) | 0.558 |
| Seroconversion of IgG antibody |  |  |  | 0.059 |
| From positive to negative | 10(13.5) | 4(26.7) | 6(10.2) | 0.110 |
| From positive to weak positive | 13(17.6) | 4(26.7) | 9(15.3) | 0.446 |
| Consistent negative | 7(9.5) | 2(13.3) | 5(8.5) | 0.624 |
| Consistent positive | 44(59.5) | 5(33.3) | 39(66.1) | **0.037** |
| Time interval of IgG antibody seroconversion, days |  |  |  |  |
| From positive to negative | 24.00(10.00-48.00) | 24.00(9.25-58.75) | 27.50(10.00-48.00) | 1.000 |
| From positive to weak positive | 7.00(4.00-22.00) | 5.50(4.25-7.75) | 11.00(4.00-30.00) | 0.279 |
| Consistent negative | 10.00(7.50-18.50) | 9.00(8.50-9.50) | 12.00(7.00-25.00) | 0.699 |
| Consistent positive | 13.50(6.75-24.00) | 13.00(12.00-14.00) | 14.00(6.00-25.00) | 0.970 |

Data are median (IQR), n (%), P values compare using $\chi^2$ test, Fisher's exact test, or Kruskal Wallis test. IQR, interquartile range.

**Figure 1.**
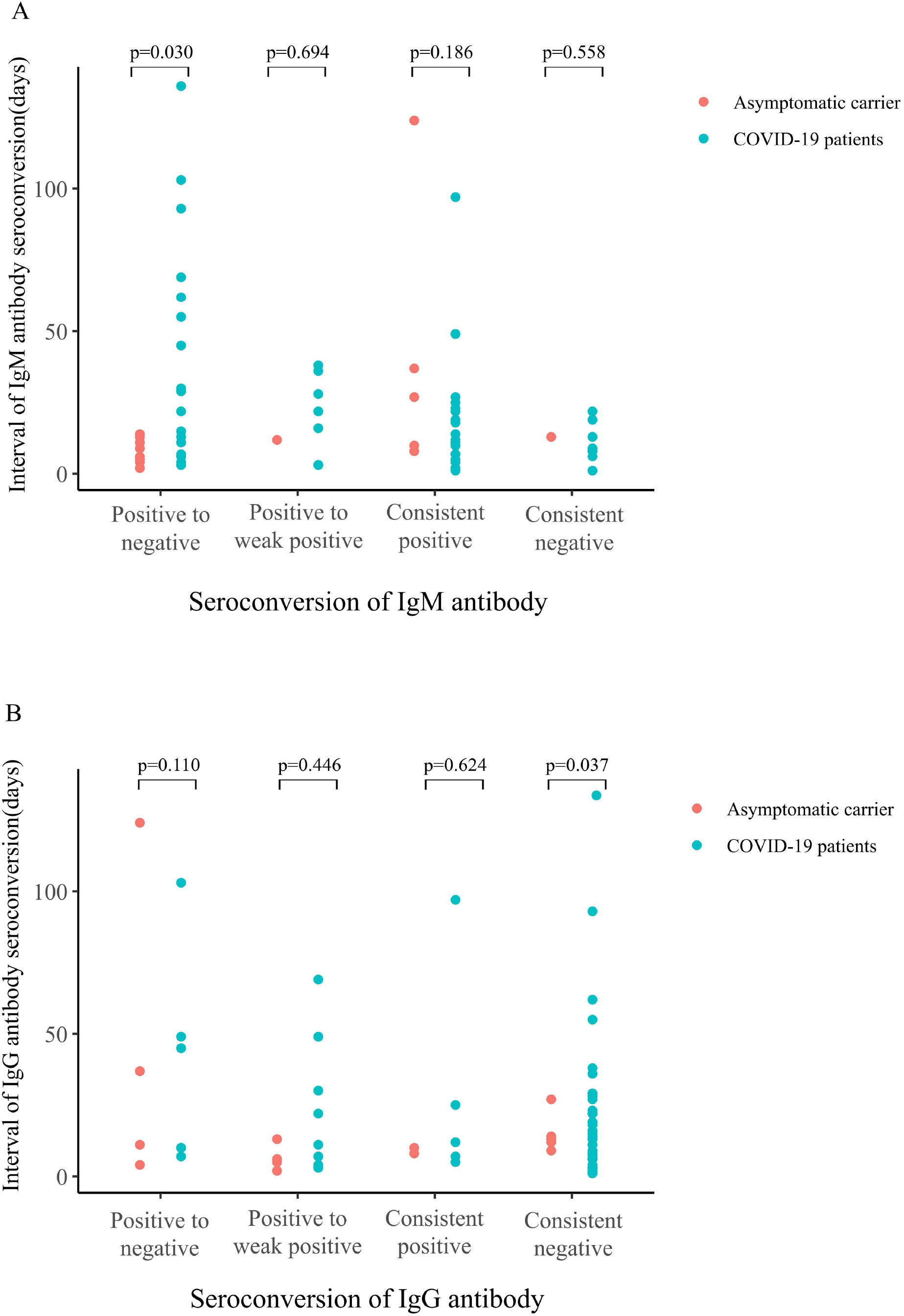
Proportion of patients owing to intra-familial transmission among asymptomatic carriers with SARS-CoV-2 infection and COVID-19 patients

### Intrafamilial transmission of SARS-CoV-2

Four family clusters were recorded in the study, 3 from Ningbo and 1 from Zhoushan. A total of 15 subjects (7 asymptomatic carriers and 8 COVID-19 patients) were confirmed to be infected by SARS-CoV-2 in the 4 familial clusters. Of the 7 asymptomatic carriers, 3 were children at the age of 12 years or younger, 3 adults aged from 18 to 60 years, and a 75-year-old woman. Of the 8 COVID-19 cases, 4 were older than 60 years and diagnosed as severe cases, 3 of the 4 severe cases had underlying diseases. The remaining 4 were mild patients at the age between 18 and 60 years. Only one of the 4 mild cases had an underlying disease. Although intra-familial transmission was the major cause of acquiring SARS-CoV-2 infection, the proportion of those who acquired SARS-CoV-2 infection via intra-familial transmission was significantly higher in asymptomatic carriers than in COVID-19 patients (89% vs. 61%, *P*=0.028) (Figure 2). In the familial cluster in Putuo, the index case’s wife who acquired the infection from his husband had a typical dynamic feature in antibodies. The titers of IgM and IgG started to decrease after nearly a month’s increase after the exposure, and then increased again (Figure 3). The second increase in IgM and IgG against SARS-CoV-2 was correlated to the time that she took care of her parents who had severe COVID-19 in the hospital.

**Figure 2.**
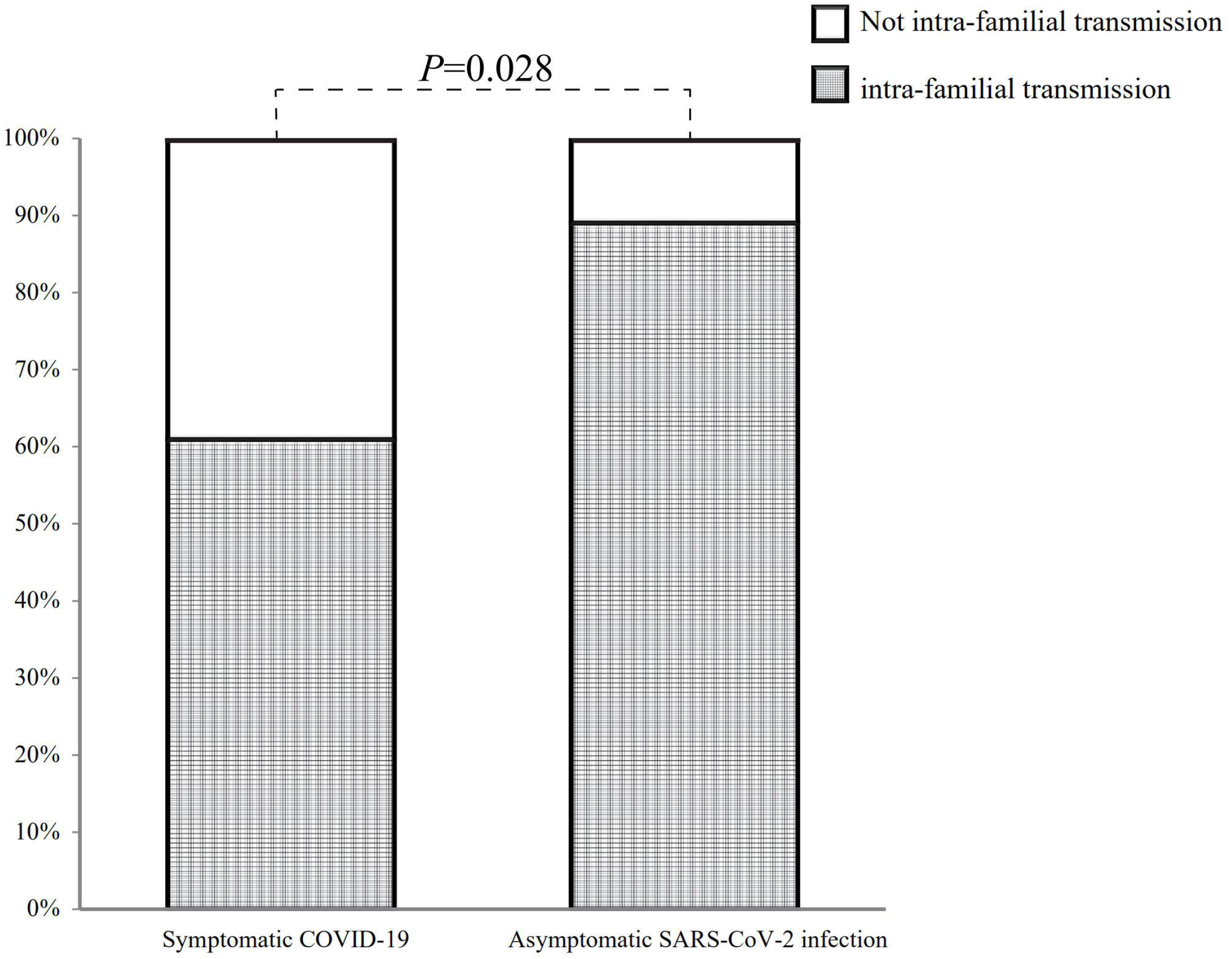
The distribution of time intervals of IgM and IgG antibody seroconversion among asymptomatic carriers with SARS-CoV-2 infection and COVID-19 patients during follow-up time

**Figure 3.**
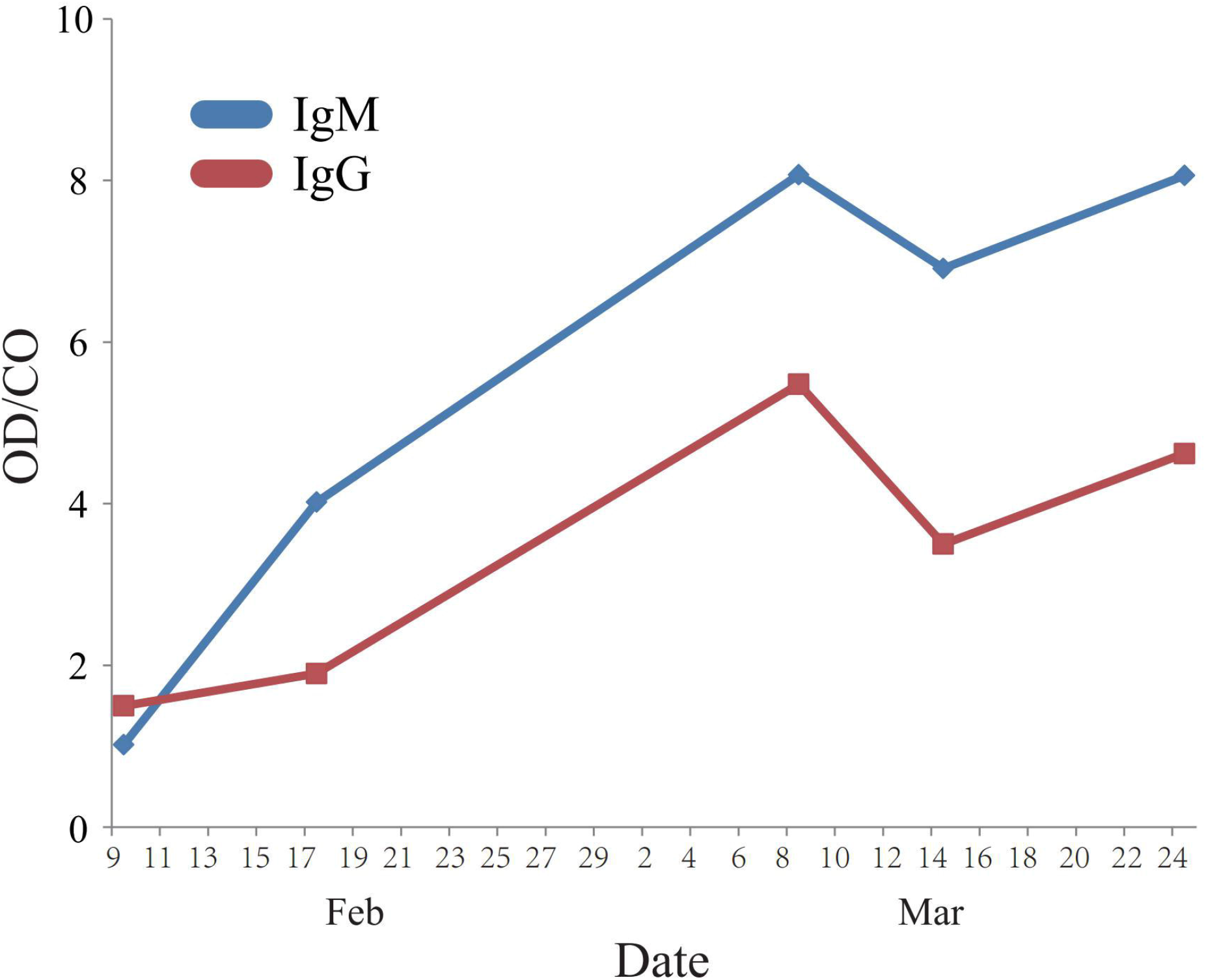
The dynamics of IgM and IgG antibody levels in a given COVID-19 patient in Putuo district of Zhoushan, Zhejiang, China, from February 9 to March 24, 2020.

## Discussion

To characterize the epidemiological features including immunological response, viral transmission, and antibody seroconversion in asymptomatic SARS-CoV-2 carriers, we made a comprehensive comparison between asymptomatic carriers and COVID-19 patients in this study. Compared to symptomatic COVID-19 patients, asymptomatic SARS-CoV-2 carriers were younger and had higher levels of circulating WBC and lymphocyte and a lower level of CRP. These data indicate that asymptomatic carriers have a stronger antiviral immunity and a lower level of systemic inflammation. It has been proven that innate and adaptive lymphocytes and inflammatory factors were closely related to disease progression of COVID-19, from mild to severe [16, 17]. In a previous prospective study, we have demonstrated that lower circulating counts of T lymphocytes, CD4^+^ T cells, and CD8^+^ T cells as well as higher circulating levels of neutrophil proportion, neutrophil/lymphocyte ratio, interleukin-6, CRP, and procalcitonin facilitate the progression of COVID-19. Of those, CD8^+^ T cell exhaustion plays an important role in the pathogenesis of COVID-19 [18]. Other studies have also shown that disease severity is negatively associated with NK cells and CD3^+^, CD4^+^, and CD8^+^ T lymphocyte levels, while intensive expansion of highly cytotoxic effector T cell subsets, such as CD4^+^ effector-granulysin, CD8^+^ effector-granulysin, and NKT CD160, is associated with convalescence of COVID-19 patients [19-21]. These evidences strongly indicate that damage of innate immunity and T cell-mediated immunity, which might be caused by proinflammatory factor-induced inflammation, play key roles in the development of COVID-19.

In this study, we also found that around 7% of asymptomatic carriers and COVID-19 patients during or after the incubation were seronegative for IgG against SARS-CoV-2, indicating SARS-CoV-2 infection might not induce sufficient humoral immunity against SARS-CoV-2. In the follow-up study, IgM seroconversion from positive to negative was much faster in asymptomatic carriers than in COVID-19 patients (*P*=0.030). The overall rate of IgG seroconversion from positive to negative or weak positive was around 30% within 160 days after the diagnosis (Table 2), indicating that IgG against SARS-CoV-2 is not stable. Virus-specific IgG decayed substantially in most individuals [22]. This was also observed in Importantly, seroconversion of IgG against SARS-CoV-2 from positive to negative or weak positive occurred 53.4% in asymptomatic carriers and 25.5% in COVID-19 patients (*P*=0.059), while consistently seropositive rate of IgG against SARS-CoV-2 was significantly higher in COVID-19 patients than in asymptomatic carriers (*P*=0.037). The similar observations concerning rapid seroconversion of the antibody against SARS-CoV-2 or short-lived immune response after mild infection were also reported in the frontline health care personnel in the US and active workers in France [23, 24]. These data indicate that humoral immunity against SARS-CoV-2 was not efficiently aroused by a relative short exposure of SARS-CoV-2 in asymptomatic carriers or in those with a stronger innate and cell immunity. Long-term seropositive rate of antibody against SARS-CoV-2 in COVID-19 patients, which has been previously reported [25, 26], indicates the feasibility of antibody-generating vaccination in the worldwide prophylactic effort. However, repeat exposure to the same virus may arouse a higher humoral immunity. A family cluster occurred in Putuo, Zhoushan should be a suitable example to address this issue (Figure 3). The index patient’s wife should be once more infected by the same SARS-CoV-2 from her patients, because both antibodies increased again after declined. Our data imply that boosting vaccination with SARS-CoV-2 might be important.

Interestingly, compared to COVID-19 patients, asymptomatic carriers had a lower level of viral load and shorter viral shedding time (Table 1). Our finding is different from a study carried out in Chongqing that asymptomatic carriers had a significantly longer duration of viral shedding than the symptomatic patients, possibly because asymptomatic carriers contained presymptomatic patients in the reported study [27]. In this study, we confirmed that the viral shedding duration was significantly longer in presymptomatic COVID-19 patients than in asymptomatic carrier (Table 1). Lower viral load and shorter viral shedding duration in asymptomatic carriers should be unlikely caused by the neutralizing antibody, because the antibody, either IgM or IgG, was declining more rapidly in asymptomatic carriers than in COVID-19 patients. Innate immunity and cell-mediated immunity should play key roles in repressing viral replication in asymptomatic carriers [28]. Lower viral load and shorter viral shedding time should be due to a relative stronger antiviral immunity, as a high viral load often predisposes adverse outcomes of COVID-19 [9, 29]. To develop effective vaccine against SARS-CoV-2, it is important to arouse the specific cell immunity, instead of focusing on humoral immunity.

In this study, we found that viral load increased from presymptomatic to symptomatic COVID-19 patients (Table 1), indicating that infectivity should be the highest at the stage of disease onset. Asymptomatic carriers had a lower level of viral load and shorter viral shedding duration, indicating that the transmissibility of asymptomatic carriers was relative weaker. In the 4 familial clusters, we found that asymptomatic carriers were mostly children and young adults, mild patients were young and middle-aged adults between 18 and 60 years, and severe cases were older than 60 years with underlying diseases. Family members were exposed to the same source of infection. However, they had diverse clinical manifestations: children were often asymptomatic whereas old members were very sick. This observation is quite in consistent with the findings of large epidemiological studies that children acquire SARS-CoV-2 infection mostly have mild respiratory symptoms or are asymptomatic, whereas elderly patients with COVID-19, especially male patients, are more likely to progress into severe-type and even die of this disease [30-33]. Thus, the host immunity and underlying inflammation, which is often affected by ageing, underlying diseases, and dysregulated macrophage response [35], should be the major determinants of disease severity of COVID-19. Although asymptomatic carriers often acquire the infection from family members, they can transmit SARS-CoV-2 into family members and hospital centers, and eventually kill aged members. As a considerable percentage of SARS-CoV-2 infections may be asymptomatic or presymptomatic, enhanced testing approaches may be needed to detect those who transmit the virus.

Our study has some limitations. First, follow-up should be extended to observe the duration of SARS-CoV-2-specific antibodies. Second, sample size of asymptomatic carriers with SARS-CoV-2 infected was relatively small.

## Conclusions

Asymptomatic carriers have a higher level of antiviral immunity and lower level of inflammation to clear SARS-CoV-2, resulting in a lower capacity of SARS-CoV-2 transmission, than do symptomatic COVID-19 patients. This antiviral immunity should not be contributable to humoral immunity because both IgM and IgG against SARS-CoV-2 are declining more rapidly in asymptomatic carriers than in COVID-19 patients. The severity of COVID-19 is associated with older age and underlying diseases in familial clustering cases. Our data also suggest that boosting vaccination with SARS-CoV-2 should be important. This study may help not only in elucidating the mechanisms by which SARS-CoV-2 interacts with host immunity in determining the outcome of SARS-CoV-2 infection, but also in optimizing the strategy for the worldwide prophylactic action to develop SARS-CoV-2 vaccine.

## Data Availability

The datasets used and analyzed during the current study are available from the corresponding author on reasonable request.

## Acknowledgments

We acknowledge all health-care workers involved in the diagnosis and treatment of patients in Ningbo and Zhoushan. We thank Ningbo CDC and Putuo CDC for providing data for patients with SARS-CoV-2.

## Ethics approval and consent to participate

This study was approved by the Ethics Commission of Ningbo CDC and Putuo CDC.

## References

1. Zhu N, et al. A Novel Coronavirus from Patients with Pneumonia in China, 2019. N Engl J Med. 2020;382:727–733. doi: 10.1056/NEJMoa2001017.

2. WHO. WHO Coronavirus Disease (COVID-19) Dashboard. https://covid19.who.int

3. Chan JF, et al. A familial cluster of pneumonia associated with the 2019 novel coronavirus indicating person-to-person transmission: a study of a family cluster. Lancet. 2020;395:514–523. doi:10.1016/S0140-6736(20)30154-9

4. Kluytmans-van den Bergh MFQ, Buiting AGM, Pas SD. Prevalence and Clinical Presentation of Health Care Workers With Symptoms of Coronavirus Disease 2019 in 2 Dutch Hospitals During an Early Phase of the Pandemic. JAMA Netw Open. 2020;3(5):e209673. doi:10.1001/jamanetworkopen.2020.9673

5. Xia XY, et al. Epidemiological and initial clinical characteristics of patients with family aggregation of COVID-19. J Clin Virol. 2020;127:104360. doi:10.1016/j.jcv.2020.104360

6. Gonzalez-Reiche AS, et al. Introductions and early spread of SARS-CoV-2 in the New York City area. Science. 2020;369:297–301. doi: 10.1126/science.abc1917.

7. Ng OT, et al. SARS-CoV-2 seroprevalence and transmission risk factors among high-risk close contacts: a retrospective cohort study. Lancet Infect Dis. 2020:S1473-3099(20)30833-1. doi: 10.1016/S1473-3099(20)30833-1.

8. Huff HV, Singh A. Asymptomatic transmission during the COVID-19 pandemic and implications for public health strategies. Clin Infect Dis. 2020:ciaa654. doi: 10.1093/cid/ciaa654.

9. Gorzalski AJ, et al. Characteristics of viral specimens collected from asymptomatic and fatal cases of COVID-19. J Biomed Res. 2020;34:431–436. doi: 10.7555/JBR.34.20200110.

10. Chen Y, et al. Epidemiological characteristics of infection in COVID-19 clo se contacts in Ningbo city. Chin J Epidemiol. 2020;41:667-671. [In Chinese]. doi: 10.3760/cma.j.cn112338-20200304-00251.

11. Li P, et al. Transmission of COVID-19 in the terminal stages of the incubation period: A familial cluster. Int J Infect Dis. 2020;96:452–453. doi: 10.1016/j.ijid.2020.03.027.

12. National Health Commission of the People’s Republic of China. Diagnosis and Treatment Protocol for COVID-19 (Trial Version 7). (updated: 2020-03-29) Available from: http://en.nhc.gov.cn/2020-03/29/c_78469.htm

13. Wang M, et al. Analytical performance evaluation of five RT-PCR kits for severe acute respiratory syndrome coronavirus 2. J Clin Lab Anal. 2020:e23643. doi: 10.1002/jcla.23643.

14. Shen C, et al. Treatment of 5 Critically Ill Patients With COVID-19 With Convalescent Plasma. JAMA. 2020;323:1582–1589. doi: 10.1001/jama.2020.4783.

15. Teng J, et al. Detection of IgM and IgG antibodies against SARS-CoV-2 in patients with autoimmune diseases. Lancet Rheumatol. 2020;2:e384–e385. doi: 10.1016/S2665-9913(20)30128-4.

16. Odak I, et al. Reappearance of effector T cells is associated with recovery from COVID-19. EBioMedicine. 2020;57:102885. doi: 10.1016/j.ebiom.2020.102885.

17. Sekine T, et al. Robust T Cell Immunity in Convalescent Individuals with Asymptomatic or Mild COVID-19. Cell. 2020;183:158-168.e14. doi: 10.1016/j.cell.2020.08.017.

18. Yang PH, et al. Increased circulating level of interleukin-6 and CD8+ T cell exhaustion are associated with progression of COVID-19. Infect Dis Poverty. 2020;9:161. doi: 10.1186/s40249-020-00780-6.

19. Chen J, et al. Clinical characteristics of asymptomatic carriers of novel cor onavirus disease 2019: A multi-center study in Jiangsu Province. Virulence. 2020;11:1557–1568. doi: 10.1080/21505594.2020.1840122.

20. Li M, et al. Elevated Exhaustion Levels of NK and CD8+ T Cells as Indicators for Progression and Prognosis of COVID-19 Disease. Front Immunol. 2020;11:580237. doi: 10.3389/fimmu.2020.580237.

21. Zhang JY, et al. Single-cell landscape of immunological responses in patients with COVID-19. Nat Immunol. 2020;21:1107–1118. doi: 10.1038/s41590-020-0762-x.

22. Chen Y, et al. Quick COVID-19 Healers Sustain Anti-SARS-CoV-2 Antibody Production. Cell. 2020;183:1496-1507.e16. doi: 10.1016/j.cell.2020.10.051.

23. Self WH, et al. Decline in SARS-CoV-2 antibodies after mild infection among frontline health care personnel in a multistate hospital network - 12 states, April-August 2020. MMWR Morb Mortal Wkly Rep. 2020;69:1762–1766. doi: 10.15585/mmwr.mm6947a2.

24. Anna F, et al. High seroprevalence but short-lived immune response to SARS-CoV-2 infection in Paris. Eur J Immunol. 2020. doi: 10.1002/eji.202049058.

25. Long QX, et al. Antibody responses to SARS-CoV-2 in patients with COVID-19. Nat Med. 2020;26:845–848. doi: 10.1038/s41591-020-0897-1.

26. Ni L, et al. Detection of SARS-CoV-2-Specific Humoral and Cellular Immunity in COVID-19 Convalescent Individuals. Immunity. 2020;52:971-977.e3. doi: 10.1016/j.immuni.2020.04.023.

27. Long QX, et al. Clinical and immunological assessment of asymptomatic SARS-CoV-2 infections. Nat Med. 2020;26:1200–1204. doi: 10.1038/s41591-020-0965-6.

28. V Vetter P, et al. Daily Viral Kinetics and Innate and Adaptive Immune Re sponse Assessment in COVID-19: a Case Series. mSphere. 2020;5:e00827-20. doi: 10.1128/mSphere.00827-20.

29. Li H, et al. High Anal Swab Viral Load Predisposes Adverse Clinical Outcomes in Severe COVID-19 Patients. Emerg Microbes Infect. 2020:1–26. doi: 10.1080/22221751.2020.1858700.

30. Castagnoli R, et al. Severe Acute Respiratory Syndrome Coronavirus 2 (SARS-CoV-2) Infection in Children and Adolescents: A Systematic Review. JAMA Pediatr. 2020;174:882–889. doi: 10.1001/jamapediatrics.2020.1467.

31. Liu K, et al. Clinical features of COVID-19 in elderly patients: A comparison with young and middle-aged patients. J Infect. 2020;80:e14–e18. doi: 10.1016/j.jinf.2020.03.005.

32. Grasselli G, et al. Baseline Characteristics and Outcomes of 1591 Patients Infected With SARS-CoV-2 Admitted to ICUs of the Lombardy Region, Italy. JAMA. 2020;323:1574–1581. doi: 10.1001/jama.2020.5394.

33. Tan YP, et al. Epidemiologic and clinical characteristics of 10 children with coronavirus disease 2019 in Changsha, China. J Clin Virol. 2020;127:104353. doi: 10.1016/j.jcv.2020.104353.

34. Mattar S, et al. Epidemiological and viral features of a cohort of SARS-Co V-2 symptomatic and asymptomatic individuals in an area of the Colombian C aribbean. Ann Clin Microbiol Antimicrob. 2020;19:58. doi: 10.1186/s12941-020-00397-5.

35. Merad M, Martin JC. Pathological inflammation in patients with COVID-19: a key role for monocytes and macrophages. Nat Rev Immunol. 2020;20:355–362. doi: 10.1038/s41577-020-0331-4.

